# Mercury exposure and health risks associated with use of skin-lightening products: A systematic review

**DOI:** 10.1101/2022.08.02.22277906

**Authors:** Ashley Bastiansz, Jessica Ewald, Verónica Rodríguez Saldaña, Andrea Santa-Rios, Niladri Basu

## Abstract

**Background:** The Minamata Convention on Mercury (Article 4) prohibits the manufacture, import or export of skin-lightening products containing mercury concentrations above 1 μg/g. However, there is a lack of knowledge surrounding the global prevalence of mercury-added skin-lightening products.

**Objective:** The objective of this study was to increase our understanding of worldwide human mercury exposure and associated health risks from the use of skin-lightening products.

**Methods:** A systematic search of peer-reviewed scientific literature was performed in four databases (PubMed, Web of Science Core Collection, Scopus, and Toxline). The initial search in July of 2018 identified 1,711 unique scientific articles, of which 34 were ultimately deemed eligible for inclusion after iterative screens at the title, abstract, and whole text levels. A second search was performed in November of 2020 using the same methods, of which another 7 scientific articles were included. All papers were organized according to four data groups 1) “Mercury in products”, 2) “Usage of products”, 3) “Human biomarkers of exposure”; and 4) “Health impacts”, prior to data extraction and synthesis.

**Results:** This review was based on data contained within 41 peer-reviewed scientific papers from 22 countries worldwide published between 2000 and 2020. In total, we captured mercury concentration values from 787 skin-lightening product samples (overall pooled central median mercury level was 0.49 μg/g, IQR: 0.02 – 5.9) and 1,042 human biomarker measurements from 863 individuals. We also synthesized usage information from 3,898 individuals, and self-reported health impacts associated with using mercury-added products from 832 individuals.

**Discussion:** This review suggests that mercury widely exists as an active ingredient in many skin-lightening products worldwide, and that users are at risk of variable, and often high exposures. These synthesized findings help increase our understanding of the health risks associated with the use of these products.

## 1. Introduction

Mercury is a global pollutant of concern to human health (Eagles-Smith et al., 2018; Ha et al., 2017). Most populations worldwide are exposed to varying degrees of mercury (Basu et al., 2018). Several forms of mercury exist naturally in the environment (i.e., metallic or elemental), inorganic and organic, and while there are differences in their toxicity, all forms can adversely impact the nervous, cardiovascular, and immune systems (Basu et al., 2018; Eagles-Smith et al., 2018; Ha et al., 2017).

Human exposures to mercury outside of occupational settings (e.g., artisanal and small-scale gold mining and dentistry) are largely realized through the consumption of mercury-contaminated seafood and contact with certain products that contain mercury (e.g., dental amalgam, pesticides, broken fluorescent lightbulbs, and batteries) (Basu et al., 2018; Eagles-Smith et al., 2018; UNEP/WHO, 2008). Another product that contains mercury is skin-lightening cosmetics, and there are increasing concerns about the dangers posed by frequent usage of such items (WHO, 2019). In certain cosmetic products, organic mercury compounds including ethylmercury, methylmercury, and phenyl mercuric salts may be used as preservatives (Al-Saleh & Al-Doush, 1997; Glahder et al., 1999; Hamann et al., 2014; Ladizinski et al., 2011). Further, inorganic mercury salts (e.g., mercurous chloride (calomel), mercuric chloride, mercurous oxide, ammoniated mercuric chloride and mercuric iodide, and ammoniated mercury), may be purposefully added into skin-lightening cosmetic products as they interfere with the tyrosinase enzyme inhibiting the skin from producing melanin, resulting in lighter skin pigmentation (Chen et al., 2020; Denton et al., 1952; Lerner, 1952). Since mercury is absorbed through the skin, mercury poisoning may arise after usage of a skin-lightening product (Olumide et al., 2008). Long-term exposure of mercury-added skin-lightening products have been associated with renal toxicity, neurological abnormalities, and dermal rashes (Chan, 2011; Palmer et al., 2000). Many women who use these products are of childbearing age, and the potential transfer of mercury from mother to fetus could have implications resulting in neurological and nephrological disorders (Bose-O’Reilly et al., 2010; Counter & Buchanan, 2004).

Skin-lightening products exist in various forms (most commonly creams and soaps), and they are used without medical supervision (Ladizinski et al., 2011). Usage of skin-lightening products is practiced worldwide, particularly in African, Asian, and Caribbean nations as well as in darker-skinned communities in Europe and North America (Dadzie & Petit, 2009; UNEP/WHO, 2008). Individuals from these communities feel motivated to use skin-lightening products to increase attractiveness, remove existing blemishes, improve skin texture, and impress their peers (Lewis et al., 2011; Osei et al., 2018; Traore et al., 2005). Societal perception of beauty continues to perpetuate the notion that lighter skin is more desirable and creates more social and professional opportunities (Peltzer et al., 2016). The market for skin-lightening products is increasing as the demographics of users continues to expand, making it one of the fastest-growing beauty industries globally. These products remain easily accessible at beauty supply stores, ethnic markets, and online (Uram et al., 2010). The illegal nature of skin-lightening products makes it difficult to differentiate the established brands since there is a strong financial motive to produce counterfeit products (Murphy et al., 2009; Prevodnik et al., 2018). As a result of the decentralized nature of these products, it is difficult to fully regulate mercury-added skin-lightening products, as countries often lack infrastructure to monitor their import and export.

The entry into force of the Minamata Convention on Mercury on 16 August 2017 was a global environmental agreement by governments set in place to reduce emissions and releases of mercury and mercury compounds to protect human health and the environment (Article 1) (UNEP, 2019). Article 4 of the Minamata Convention mandated that Parties must ban the manufacture, import, and export of products that include creams and soaps with a mercury content higher than 1 μg/g after 2020 (UNEP, 2019). This value of mercury has already formed a basis for regulatory action by organizations such as the US Food and Drug Administration (FDA) and Health Canada (Government of Canada, 2012; U.S. FDA, 2020). Despite such limits (and similar ones set by many governments worldwide), many mercury-added skin-lightening products have mercury levels that exceed 1 μg/g (Hamann et al., 2014). Issues detailed in the previous paragraphs, coupled with our lack of knowledge concerning the global prevalence of these products and their mercury content, pose challenges for scientists, regulators, and manufacturers. Accordingly, the overall objective of this study was to increase our understanding of worldwide human mercury exposure and associated health risks from the use of skin-lightening products. This objective was realized through a systematic review of the peer-reviewed scientific literature centered on the following four data groups: 1) “Mercury in products” for the characterization of mercury in products (e.g., concentration, speciation, types of products); 2) “Usage of products”, for users of skin-lightening products (e.g., socioeconomic variables, demographics, exposure variables); 3) “Human biomarkers of exposure” for human biomarkers of skin-lightening users (e.g., hair, urine, blood); and 4) “Health impacts” for associated health outcomes of usage.

## 2. Methods

### 2.1 Search strategy

A protocol for the search strategy was developed in July 2018 and then revisited in November 2020. We reviewed key resources [Preferred reporting items for systematic reviews and meta-analyses (PRISMA); http://www.prisma-statement.org/; (OHAT, 2015)] including a systematic review concerning methylmercury exposure from seafood consumption (Sheehan et al., 2014) and a recent review concerning worldwide mercury exposures as part of the 2018 UN Global Mercury Assessment (Basu et al., 2018). The protocol defined study selection criteria and the type of data that would be extracted for each study and was uploaded to PROSPERO on April 28, 2020 (CRD42020160299).

A systematic search of peer-reviewed scientific literature was used to identify relevant studies. An electronic search was performed in four databases (PubMed, Web of Science Core Collection, Scopus, and Toxline), with the last search carried out on 20 November 2020 as outlined in the PRISMA flow diagram (Figure 1). The search strategy included two Boolean search phrases that were combined with AND: #1 – mercury OR Hg OR MeHg OR *Mercur*; and #2 - cosmetic* OR lotion* OR cream* OR soap* OR shampoo* OR makeup OR whitening OR lightening OR bleaching OR mascara. In addition to the systematic search, we considered grey literature and references cited in read works. We focused on works that were written in English, French, Spanish, and Portuguese.

**Figure 1.**
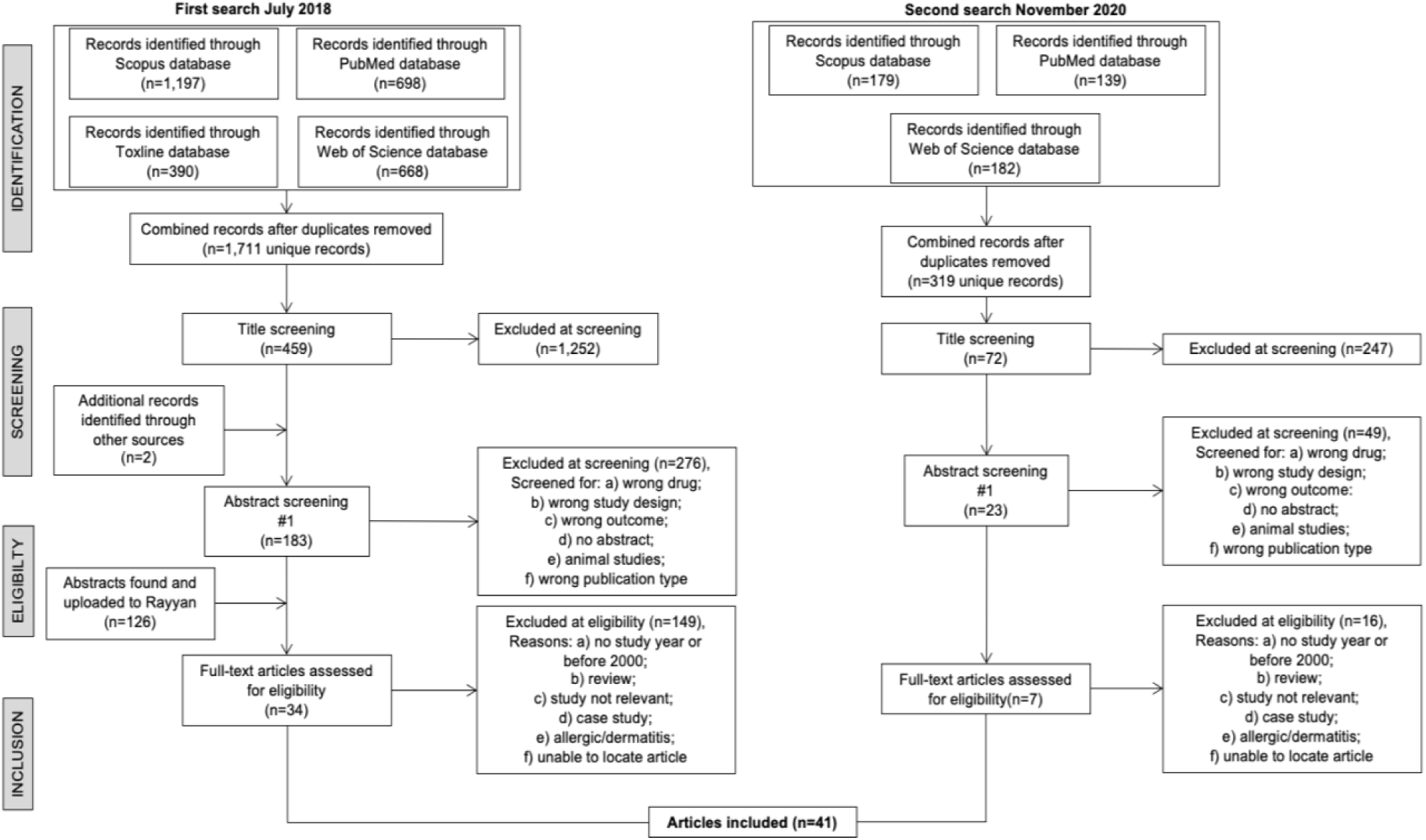
PRISMA flow diagram detailing how articles were identified, reviewed, and deemed eligible for inclusion in the current study.

### 2.2 Study selection criteria

Scientific papers were identified and reviewed in July 2018, and then again in November 2020. We restricted studies to those published in or after 2000 and only original articles were included for analysis. After the initial literature search (July 2018), duplicates were removed, and a list of 1,711 unique references was imported to Mendeley. Next, the publication titles were reviewed to determine their relevance which resulted in the inclusion of 459 unique citations. Publications excluded at this stage were labelled with one of the following primary reasons: “No abstract”, “Background paper”, “Foreign language”, “Wrong drug”, “Wrong outcome”; “Wrong population”, “Wrong population type”, “Wrong study design”, and “Wrong study duration”. Medical case reports were excluded from the main study but were tagged so that they could be qualitatively summarized separately from the formal systematic review. This screening activity was carried out by dividing the total number of publications into two parts, and each part was screened by three reviewers (J.E., V.S. and A.S.) to avoid conflicts. This effort resulted in the inclusion of 183 publications and the exclusion of 276.

A second literature search, using the previously described methods, was conducted in November 2020 using three databases (PubMed, Web of Science Core Collection, and Scopus) to identify additional scientific literature that had been published after the first literature search. After removing duplicates, 319 publications were identified for screening. These articles were assessed in the same manner as the first search, and from this activity we identified 23 additional publications for inclusion.

From the first and second literature searches, 183 and 23 publications, respectively, were included for full text screening. At this point, we focused on binning publications (in particular, the studies’ data and information) into four groups: 1) “Mercury in products” for the characterization of mercury in products (e.g., concentration, speciation, types of products); 2) “Usage of products”, for users of skin-lightening products (e.g., socioeconomic variables, demographics, exposure variables); 3) “Human biomarkers of exposure” for human biomarkers (e.g., mercury measures in hair, urine, or blood) of skin-lightening users; and 4) “Health impacts” for associated health outcomes of usage. Based on initial reviews of the available data, we decided to focus the first (“Mercury in products”) and third (“Human biomarkers of exposure”) groups on papers in which quantitative data could be extracted, and papers from the second (“Usage of products”) and fourth (“Health impacts”) groups on papers in which data could be explored in a semi-quantitative manner. Two studies external from the literature search (Kinabo, 2005; Voegborlo et al., 2008) identified through an included publication (Agorku et al., 2016) were also included for analysis. After carrying out the full text screening, 34 and 7 papers from the first and second search, respectively, were identified for the data extraction phase, thus resulting in a total of 41 papers.

### 2.3 Data Extraction and Analysis

To summarize mercury concentrations in skin-lightening product data and human biomarkers, the approach of Sheehan et al. (2014) was followed. Briefly, we reported on two summary distributions [central (median) and upper] for each skin-lightening product by pooling the respective data together over relevant studies. When studies had multiple measurements for central exposures, median measurements were favored over the mean. Throughout this report, we refer to total mercury as “mercury” measurements in a given skin-lightening product or human biomarker sample type.

#### 2.3.1 “Mercury in products” Data Group

Studies were organized by location (country, WHO region) of product manufacture and purchase, and categorized into type of product (cream, soap, facial soap, other), and method of purchase (in store, online). Products categorized as “other” included scrubs, balms, oils, pills, personal mixtures as well as products that were not specifically labelled. Personal mixtures of multiple products and at home remedies are commonly used to increase potency and whitening effect. From each study, data was extracted on sample size (number) and volume of sample (mL or g), along with key information on the mercury measurement including the inorganic (ammoniated mercury) and organic mercury compounds (phenyl mercuric salts, thiomersal), testing instrumentation [cold vapor-atomic absorption spectrophotometry (CVAAS), X-ray fluorescence (XRF), inductively coupled plasma mass spectrometry (ICP-MS), etc.], analytical quality control (reported detection limit, accuracy, precision), and a measure of the central mercury concentration values (mean or median, as μg/g). Skin-lightening cosmetic samples assessed using XRF instrumentation and commercial screening kits were excluded from the analysis due to concerns surrounding the reliability of the measures they report. Products manufactured in the same geographic location (country, WHO region) and of the same product type were collectively grouped together. We considered mercury as an active ingredient in samples with concentrations greater than 1 μg/g. Non-detectable mercury concentration values were entered as half the detection limit as recommended in the US EPA’s Regional Guidance on Handling Chemical Concentration Data Near the Detection Limit in Risk Assessment (Smith, 1991).

#### 2.3.2 “Usage of products” Data Group

A review publication by Sagoe et al. (2019) was identified and adopted as a framework for this data group. After removing duplicate studies from our literature search as well as studies published before 2000, a total of 16 studies were included for analysis. Since usage patterns are irrespective of product ingredients (i.e., mercury, hydroquinone, topical corticosteroids), usage patterns of all skin-lightening product users were considered. We extracted information on study type, type of bleaching products, and demographics of users (i.e., location, sex, age, marital status, occupation, education level, average monthly income), and key factors surrounding exposure characteristics were evaluated: (1) application (i.e., face, whole body), (2) frequency of application (per day), (3) quantity used (grams per month); and (4) duration (months).

#### 2.3.3 “Human biomarkers of exposure” Data Group

For biomarkers (hair, urine, blood) of mercury exposure, we extracted data on the populations’ characteristics (age, sex, sample size) and location (country, WHO region), as well as mercury exposure measurements [mercury speciation, testing instrumentation (CVAAS, XRF, ICP-MS, etc.), analytical quality control (reported detection limit, accuracy, precision, testing instrumentation)], and a measure of central tendency (mean or median).

#### 2.3.4 “Health impacts” Data Group

Reported health impacts associated with the use of mercury-added skin-lightening products were reviewed in a semi-quantitative manner. Key metadata (e.g., location, type of study, age, sample size, sex) and measurement information (level of mercury in biomarker, associated health effect) were recorded as available. Individuals reporting health impacts in combination with other mercury containing products or skin-lightening products not containing mercury were not included. We focused this work only on current users of skin-lightening products.

### 2.4 Quality Assessment/Risk of Bias

We assessed risk of bias and study quality specifically for the “Mercury in products” and “Human biomarkers of exposure” data groups. This was performed using the framework published by Hu et al. (2018) and the National Toxicology Program’s OHAT framework (OHAT, 2015). Study quality was assessed by nine items that were grouped into three risk of bias categories: (1) mercury exposure detection bias [i.e., measurement instrument, use of reference material, replicate measures, acceptable limit of detection (LOD)]; (2) selection bias (i.e., selection method, sample size, mercury exposure characteristics); and (3) other biases (i.e., demographics, key descriptive measures). Each of the nine items was scored “0” (high risk of bias), “1” (moderate risk of bias) or “2” (low risk of bias). Author (A.B.) independently scored each paper and consulted with other authors (J. E., A. S., and V. S.) in cases where additional consensus was required. For the skin-lightening product sample sizes, the sample size item was given a score of “0” if studies assessed <10 products, “1” for 10-30 products, and “2” for >30 products. For biomarker samples, the sample size item was given a score of “0” if studies assessed <50 biomarker samples, “1” for 50-200 biomarker samples, and “2” for >200 biomarker samples. The overall study rating was based on an assessment of the nine items. Total scores ranged between 0-18 and characterized as: high risk/low quality (0-5), moderate risk/quality (6-12), and high quality/low risk (13-18). Studies characterized as “high risk/low quality” were further reviewed before being deemed appropriate for inclusion.

## 3. Results

### 3.1 Overview of studies

This review was based on data contained within 41 peer-reviewed scientific papers from 22 countries published between 2000 and 2020. In total, we captured mercury concentration values from 787 skin-lightening product samples, and 1,042 biomarker measurements from 863 individuals. The average sample size across “Mercury in products” studies was 36 products with a range of 2 to 191 skin-lightening products reported upon in each study. The average sample size for the “Human biomarkers of exposure” group excluding non-users (i.e., family members, control groups) was 88 biomarker samples with a range between 11 to 282 biomarker samples per study. In addition, we compiled usage information from 3,898 individuals, and self-reported health impacts associated with using mercury-added products from 832 individuals.

Studies from the two quantitative data groups (i.e., mercury levels in products, and human biomonitoring studies) were largely situated, according to WHO region locations, in the Eastern-Mediterranean (27.6%), Western Pacific (24.1%), the Americas (24.1%), and Africa (17.2%) regions, with fewer studies in South-East Asia (3.4%) and Europe (3.4%) (Table 2). Studies concerning usage of products and health impacts were largely situated in Africa (31.3%) and the Americas (31.3%), Eastern-Mediterranean (18.8%), Western-Pacific (12.5%), with the fewest in South-East Asia (6.3%) and none from Europe.

**Table 1.** Overview of studies included in this review.

| Data group | No. of studies | Study analysis type | Summary |
| --- | --- | --- | --- |
| “Mercury in products” | 25 | Quantitative | Studies analyzing mercury concentrations in skin-lightening cosmetic samples were collected from 16 countries/territories with sample sizes ranging from 2 and 191 per study |
| “Usage of products” | 16 | Semi-quantitative | Studies were collected from nine countries to describe skin-lightening usage patterns (e.g., frequency, duration, application, quantity) amongst users |
| “Human biomarkers of exposure” | 9 | Quantitative | Studies were collected from six countries/territories analyzing mercury concentrations in blood, urine, and hair with sample sizes ranging between 11 and 465 samples per study |
| “Health impacts” | 5 | Semi-quantitative | Studies were collected from four countries/territories observing associated health effects from mercury-added skin-lightening products |

**Table 2.** Count of studies and their distribution across WHO regions, along with the total number of items (for “Hg in products” and “Human biomarkers of exposure” groups) or individuals (for “Usage of products” and “Health impacts” groups) included in this review.

| **Data Group** | **No. of studies** | **No. of Items or Individuals** | **Africa** | **Americas** | **Eastern Mediterranean** | **Europe** | **South-East Asia** | **Western Pacific** |
| --- | --- | --- | --- | --- | --- | --- | --- | --- |
| 1) “Mercury in products” | 25 | 787 | 5 | 5 | 7 | 1 | 1 | 6 |
| 2) “Usage of products” | 16 | 3,898 | 5 | 3 | 3 | 0 | 1 | 1 |
| 3) “Human biomarkers of exposure” | 9 | 1,042 | 1 | 3 | 1 | 0 | 1 | 3 |
| 4) “Health impacts” | 5 | 832 | 0 | 3 | 0 | 0 | 0 | 2 |

### 3.2 Quality Assessment

There were 25 studies grouped under “Mercury in products”, and these studies had a mean total risk of bias score of 10.3 with individual total scores ranging from 4 to 14. A total of three (11.1%) studies were categorized as high quality/low risk of bias, whereas 23 (85.2%) studies met the moderate quality/risk of bias, and one (3.7%) study was considered low quality/low risk of bias (Table 3). There were nine studies grouped under “Human biomarkers of exposure”, and these studies had a mean total risk of bias score of 9.6 with individual total scores ranging from 6 to 12. All these studies met the moderate quality/risk of bias. We note that all studies used convenience sampling methods to retrieve human biomarker and skin-lightening product samples.

**Table 3.** Study quality scores for nine items that assess risk of bias within studies of the 1) “Mercury in products” and 3) “Human biomarkers of exposure” data groups. Each of the nine items were given a score of either 0, 1, or 2, and summed for an overall study quality score [0 to 5 (low quality/high risk of bias), 6 to 12 (moderate quality/risk of bias), and 13 to 18 (high quality/low risk of bias)]. The nine items in the top row correspond to 1) measurement instrument reported, 2) accuracy through use of reference material, 3) precision through use of replicate measures, 4) LOD reported and acceptable, 5) selection method (convenience/random), 6) sample size, 7) demographics reported, 8) Hg exposure characteristics reported; and 9) paper had key descriptive measures reported. *Short communication study, and methods for testing products were not clearly stated.

| Data Group | Reference | 1 | 2 | 3 | 4 | 5 | 6 | 7 | 8 | 9 | Total Score | Overall |
| --- | --- | --- | --- | --- | --- | --- | --- | --- | --- | --- | --- | --- |
| Human Biomarkers | (Sin & Tsang, 2003) | 0 | 0 | 0 | 0 | 1 | 2 | 2 | 2 | 2 | 9 | Moderate |
|  | (Weldon et al., 2000) | 0 | 0 | 0 | 0 | 1 | 2 | 2 | 2 | 2 | 9 | Moderate |
|  | (Fakour & Esmaili-Sari, 2014) | 2 | 2 | 2 | 0 | 2 | 0 | 2 | 0 | 1 | 11 | Moderate |
|  | (McKelvey et al., 2011) | 2 | 2 | 2 | 2 | 1 | 0 | 2 | 0 | 1 | 12 | Moderate |
|  | *(Harada et al., 2001) | 0 | 2 | 2 | 0 | 1 | 0 | 2 | 2 | 1 | 10 | Moderate |
|  | (McRill et al., 2000) | 0 | 0 | 0 | 0 | 1 | 1 | 2 | 0 | 2 | 6 | Moderate |
|  | (Abbas et al., 2020) | 2 | 0 | 0 | 1 | 0 | 1 | 2 | 2 | 2 | 10 | Moderate |
|  | (Kanwal et al., 2019) | 2 | 2 | 0 | 2 | 0 | 0 | 1 | 0 | 2 | 9 | Moderate |
|  | (Kinabo, 2005) | 2 | 0 | 0 | 2 | 0 | 0 | 2 | 2 | 2 | 10 | Moderate |
| Mercury in products | (Agorku et al., 2016) | 2 | 2 | 2 | 2 | 1 | 2 | 0 | 0 | 1 | 12 | Moderate |
|  | (Murphy et al., 2009) | 2 | 2 | 2 | 1 | 1 | 1 | 0 | 0 | 1 | 10 | Moderate |
|  | (Ho et al., 2017) | 2 | 2 | 2 | 2 | 1 | 1 | 0 | 0 | 2 | 12 | Moderate |
|  | *(Harada et al., 2001) | 0 | 0 | 0 | 0 | 0 | 0 | 2 | 1 | 1 | 4 | Low |
|  | (Zainy, 2015) | 2 | 2 | 2 | 1 | 0 | 1 | 0 | 0 | 1 | 9 | Moderate |
|  | (Peregrino et al., 2011) | 2 | 2 | 2 | 2 | 0 | 2 | 0 | 0 | 1 | 11 | Moderate |
|  | (Gbeto & Amyot, 2016) | 2 | 2 | 2 | 2 | 1 | 2 | 0 | 0 | 2 | 13 | Good |
|  | (Al-Saleh et al., 2012) | 2 | 2 | 2 | 2 | 0 | 2 | 0 | 0 | 2 | 12 | Moderate |
|  | (Yang et al., 2014) | 2 | 2 | 2 | 2 | 0 | 0 | 0 | 0 | 1 | 9 | Moderate |
|  | (Salama, 2016) | 2 | 2 | 2 | 1 | 0 | 0 | 0 | 0 | 0 | 7 | Moderate |
|  | (Wang & Zhang, 2015) | 2 | 2 | 2 | 2 | 0 | 1 | 0 | 0 | 0 | 9 | Moderate |
|  | (McKelvey et al., 2011) | 0 | 0 | 0 | 0 | 1 | 1 | 2 | 0 | 2 | 6 | Moderate |
|  | (Ashraf et al., 2020) | 2 | 2 | 2 | 1 | 0 | 1 | 0 | 0 | 1 | 9 | Moderate |
|  | (Selvaraju et al., 2020) | 2 | 2 | 2 | 2 | 1 | 1 | 0 | 0 | 2 | 12 | Moderate |
|  | (Abbas et al., 2020) | 2 | 0 | 0 | 1 | 0 | 1 | 2 | 2 | 2 | 10 | Moderate |
|  | (Kanwal et al., 2019) | 2 | 2 | 0 | 2 | 0 | 0 | 0 | 0 | 2 | 8 | Moderate |
|  | (Kinabo, 2005) | 2 | 0 | 0 | 2 | 0 | 2 | 2 | 2 | 2 | 12 | Moderate |
|  | (Voegborlo et al., 2008) | 2 | 2 | 2 | 1 | 1 | 2 | 0 | 0 | 2 | 12 | Moderate |
|  | (Maneli et al., 2016) | 2 | 2 | 2 | 1 | 0 | 1 | 0 | 0 | 2 | 10 | Moderate |
|  | (Alqadami et al., 2017) | 2 | 2 | 2 | 2 | 0 | 1 | 0 | 0 | 2 | 11 | Moderate |
|  | (Alqadami et al., 2013) | 2 | 2 | 2 | 2 | 0 | 2 | 0 | 0 | 2 | 12 | Moderate |
|  | (Ababneh & Al-Momani, 2018) | 2 | 2 | 2 | 2 | 1 | 2 | 0 | 0 | 1 | 12 | Moderate |
|  | (Mohammed et al., 2017) | 2 | 2 | 2 | 2 | 0 | 1 | 0 | 0 | 1 | 10 | Moderate |
|  | (Amponsah et al., 2014) | 2 | 2 | 2 | 1 | 1 | 2 | 0 | 0 | 2 | 12 | Moderate |
|  | (Cristaudo et al., 2013) | 2 | 1 | 2 | 2 | 0 | 1 | 2 | 2 | 2 | 14 | Good |

In terms of exposure detection bias, the most commonly used mercury analyzer for skin-lightening products was cold vapor atomic absorption spectrometry (CVAAS) (59.2%), followed by atomic absorption spectroscopy (AAS) (11.9%), and inductively coupled plasma-optical emission spectrometry (ICP-OES) (9.9%) (Table S1). For human biomarker measurements, the most commonly used instrument was particle induced X-ray emission (PIXE) (14.2%), followed by advanced mercury analyzer (AMA) (5.6%), and AAS (3.8%) (Table S2). Notably, a detection instrument was not listed for 768 (73.7%) of the biomarker measurements. Most “Mercury in products” (84%) and “Human biomarkers of exposure” (80%) studies considered analytical quality control measures by fully reporting on three of the four key items (measurement instrument, accuracy, precision, and detection limit). In total, eight quantitative studies reported no quality control measures since product and human biomarker samples in these studies were sent to external laboratories for analysis. As a result, these studies were carefully examined and deemed appropriate for inclusion. In terms of selection bias, all skin-lightening products and biomarker samples were conveniently sampled from stores and markets, and individuals, respectively.

For the “Mercury in products” data group, four studies (16%) tested <10 products, 11 studies (44%) tested between 10-30 products, and 10 studies (40%) tested >30 products. For the “Human biomarkers of exposure” data group, five studies tested <50 biomarker samples, two studies (22.2%) tested between 50 and 200 biomarkers, and two studies (22.2%) tested >200 biomarker samples. User demographics (i.e., age, sex, location) were reported upon in all the “Human biomarkers of exposure” studies, however most (84%) “Mercury in products” studies did not report on user demographics. Mercury exposure characteristics were only reported in four (16%) “Mercury in products” studies and in five (55.6%) “Human biomarkers of exposure” studies. In terms of other biases, only three (12%) “Mercury in products” studies did not list any descriptive measures (i.e., mean, upper values), whereas all “Human biomarkers of exposure” studies listed descriptive measures.

### 3.3 “Mercury in products” Data Group

In the analysis of skin-lightening products, we grouped products according to WHO regions of purchase and manufacture, product type, and mercury product concentration. A total of 787 skin-lightening products were identified from 25 studies which included creams (70.1%), soaps (19.3%), facial cream (4.6%), and other products (6.0%) (Excel Table S1). The overall pooled central median mercury concentration in skin-lightening products was 0.49 μg/g (IQR: 0.02 and 5.9 μg/g) (Figure 2). In Figure 2 we excluded 49 skin-lightening products since mercury concentrations were only available as ranges and not discrete values. Mercury was an active ingredient (mercury concentration >1 μg/g) in 196 (24.9%) products. The mercury concentration varied across product types and geographic regions. While the average values of mercury in the various products were similar, the maximum reported value for creams was 314,387 μg/g, followed by facial cream (35,824 μg/g), soap (8,665 μg/g) and other products (2,700 μg/g).

**Figure 2.**
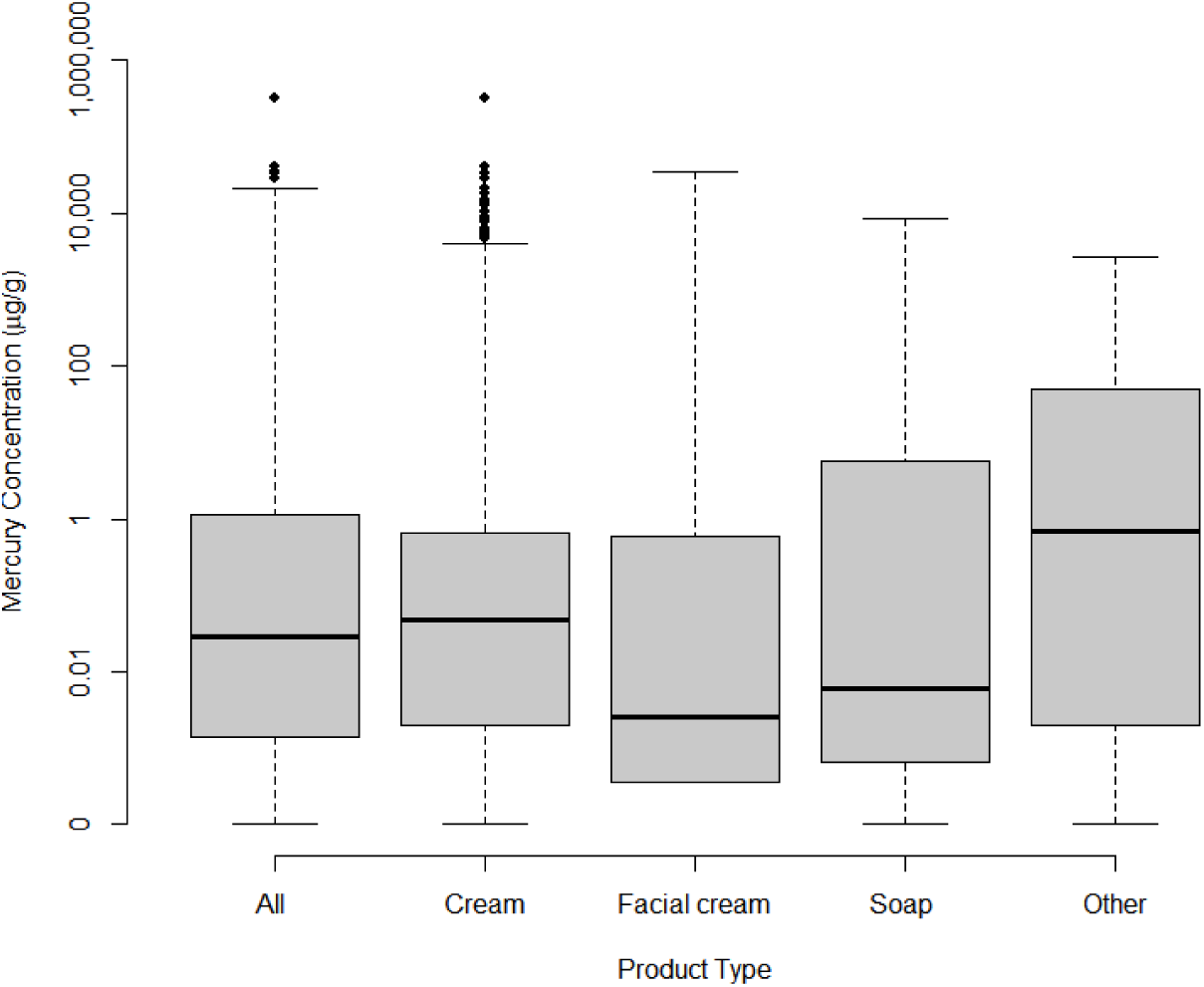
Boxplots showing mercury concentrations (μg Hg/g) found in all 738 skin-lightening products (as well as in creams, facial creams, soaps, and other categorized items).

Most skin-lightening products were purchased in store (95.3%), with the procurement of the rest not reported by authors or mislabeled. None of the products reported the presence of phenyl mercuric salts or thiomersal, whereas 0.4% of creams reported containing ammoniated mercury. Skin-lightening products were manufactured in 33 countries mainly within Europe (22.6%), South-East Asia (8.4%), and African (7.8%) regions, versus those in the Americas (6.5%), Western-Pacific (2.9%), and Eastern-Mediterranean (2.9%) regions (Figure 3). The manufacture location was unknown or unlisted for nearly half (48.9%) of the products. Products were purchased in 20 countries within the African (55%), Eastern-Mediterranean (20.5%), American (9.9%), Western-Pacific (9.4%), South-East Asian (3.4%) and Europe (1.8%) regions (Figure 4). Despite Europe manufacturing the largest quantity of products respective to the other regions, only 1.8% of skin-lightening were purchased in Europe.

**Figure 3.**
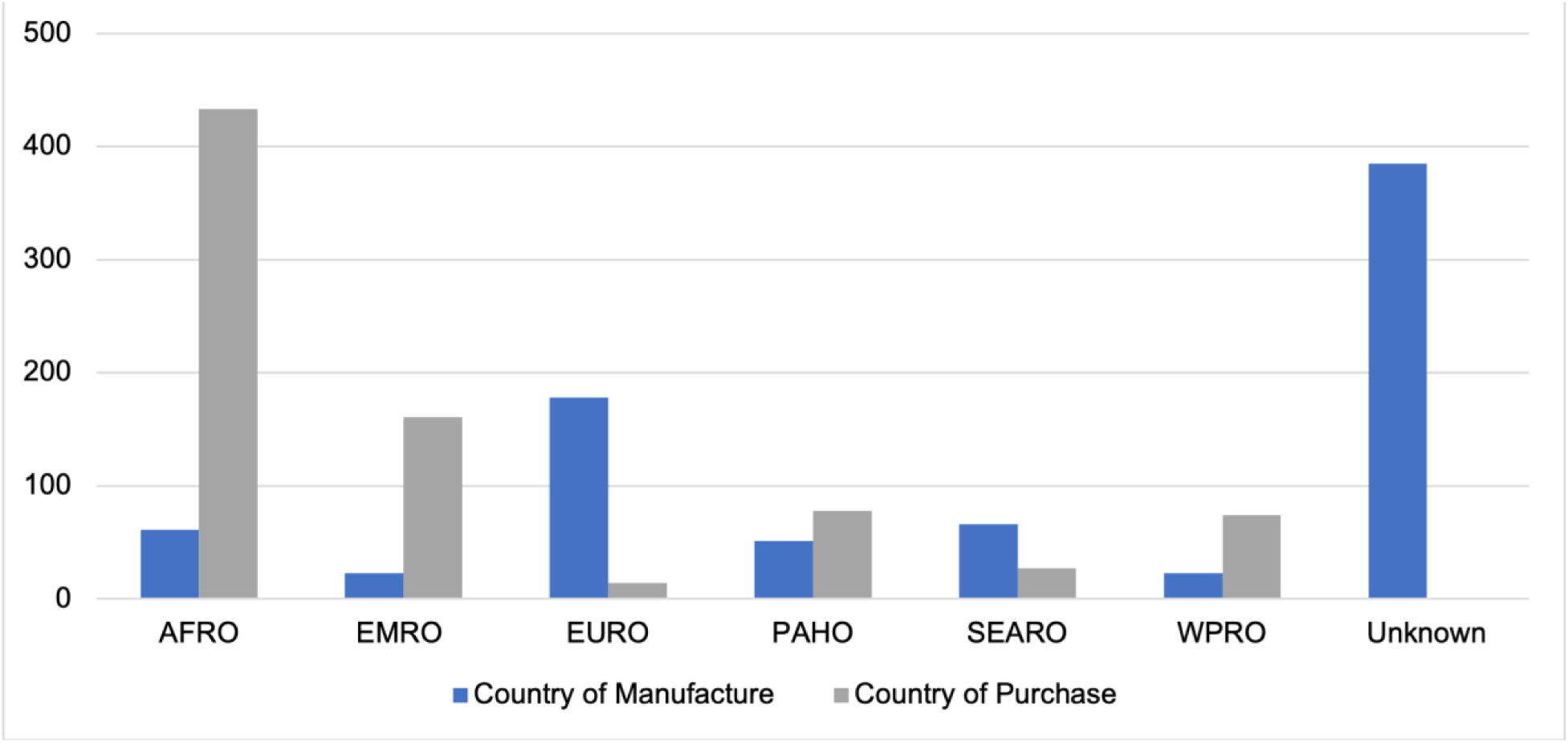
Bar plots showing quantity and types of 787 skin-lightening products manufactured and purchased according to the WHO regions.

**Figure 4.**
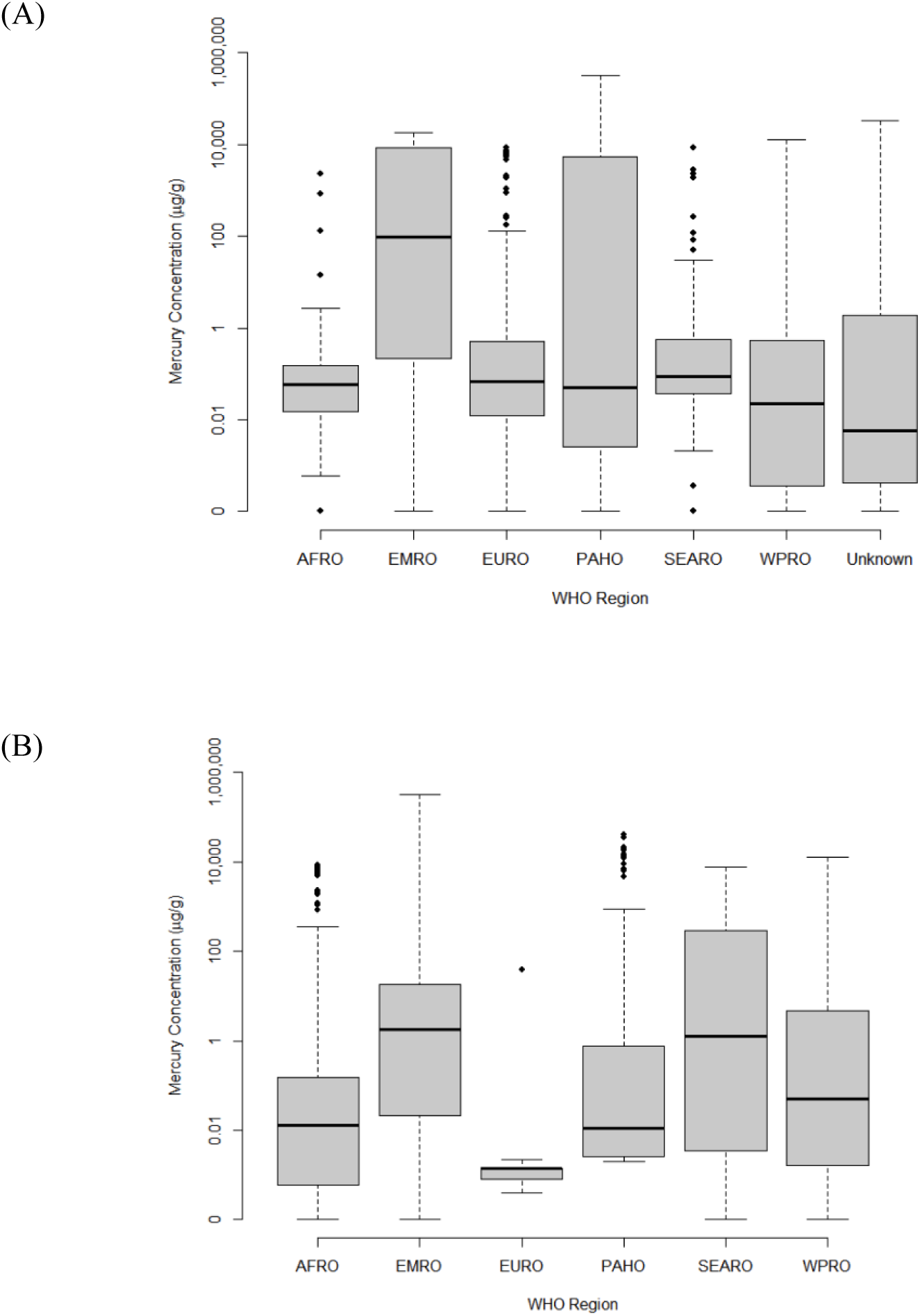
Boxplots showing mercury concentrations (μg Hg/g) in 738 skin-lightening products (creams, facial creams, soaps, other) across six WHO regions where products were (A) manufactured and (B) purchased.

### 3.4 “Usage of products” Data Group

A total of 3,898 individuals surveyed from 16 studies were included in the “Usage of products” data group (Excel Table S2). Not all the individuals were surveyed on all four key exposure characteristics (e.g., application, frequency, duration, quantity), and thus there are not 3,898 data points for each of the exposure characteristics. In terms of application, a total of 1,708 individuals from seven studies reported on application areas (e.g., face, body). Users were most likely to apply skin-lightening products to their face (53.1%) in comparison to “other” application areas (e.g., hands, neck, non-exposed sun areas) (22.4%) and whole body (18.6%). In one study, the sum of percentages exceeds 100% which may be attributed to individuals reporting on more than one application area (Alghamdi, 2010). In terms of frequency, we reported on information from a total of 2,525 individuals from 10 studies. From this data, most individuals were using products less than one time to three times per day (87.2%), more than three times per day (0.8%) and other unspecified times (8.6%). In terms of duration, we collected information from a total of 2,098 individuals from 10 studies. The duration of usage of skin-lightening products ranged between 1 month to 30 years. Out of 1,187 individuals surveyed from five studies, a total of 385 (32.4%) individuals used skin-lightening products from less than a year, 323 (27.2%) individuals used products between 1 to 5 years, 133 (11.2%) individuals used products between 6 to 10 years, 168 (14.2%) individuals used products between 11 to 20 years, 100 (8.4%) individuals used products above 20 years and 78 (6.6%) individuals used it for an unknown period of time (Alanzi et al., 2018; Brinton et al., 2018; Harada et al., 2001; Kinabo, 2005; Sendrasoa et al., 2017). A total of 4 studies reported on 1,008 individuals that used skin-lightening products for a duration of 2 months to 30 years, and a total of 157 and 105 individuals from two studies reported a mean duration period of between 20 and 26 months, respectively (Abbas et al., 2020; Alghamdi, 2010; Atadokpédé et al., 2015; Lartey et al., 2017; Ly et al., 2007). In terms of quantity, we collected information from 1,015 individuals from four studies on the number of skin-lightening products used per month. From a total of 335 individuals, 94 (28%) individuals used less than and up to 10 g of product per month, 160 (47.8%) individuals used between 11 to 50 g of product per month, and 81 (24.2%) individuals used above 50 g of product per month (Abbas et al., 2020; Alanzi et al., 2018). In two studies, 703 (58.7%) individuals used between 2 to 600 g of product per month with 194 (16.2%) using a median of 95 g of product per month and 509 (42.5%) individuals using a median of 90 g of product per month (Alghamdi, 2010; Mahe et al., 2003).

### 3.5 “Human biomarkers of exposure” Data Group

A total of 1,042 biomarker samples (urine – 56.3%, hair – 26.1%, blood – 17.6%) were collected from 863 skin-lightening product users, family members of users, and control groups between 14 and 79 years of age (Table 4). Only three studies (Abbas et al., 2020; Kinabo, 2005; McRill et al., 2000) included control groups (e.g., non-users, family members of users) to assess and compare biomarker concentrations.

**Table 4.**
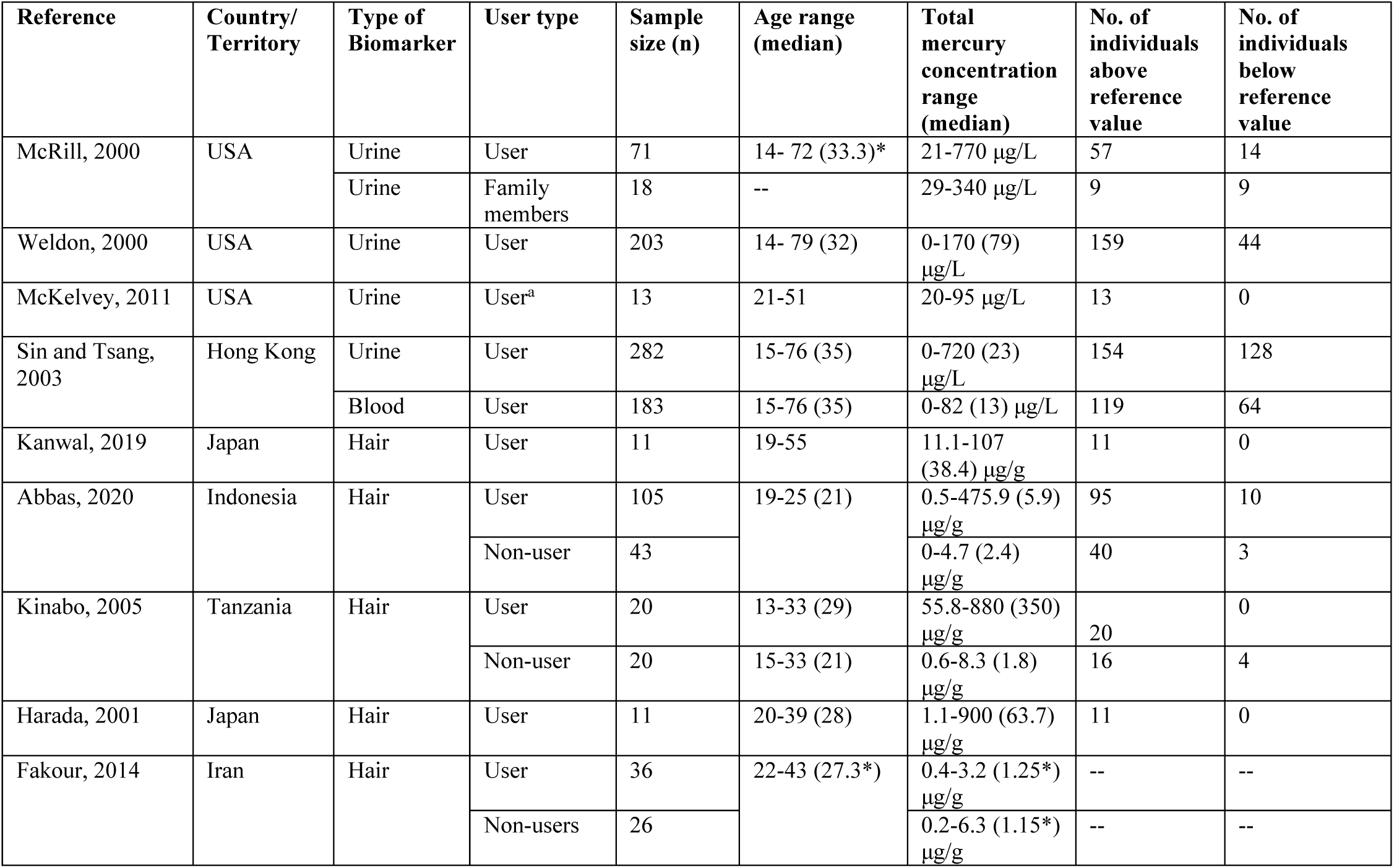
Summary of mercury concentrations from the “Human Biomarkers of Exposure” data group. The upper reference values were: urine (20 μg/L), blood (10 μg/L), hair (1 μg/g). ^a^ Includes nine known product users, and four individuals suspected of using products. ^b^ Control group includes concentrations from both users and non-users. *Mean values used since median values were not available.

#### 3.5.1 Urine mercury levels

Total mercury levels in urine across 569 skin-lightening product users from Hong Kong and US-Mexican border states (Arizona, California, New Mexico, and Texas) ranged between 0 and 770 μg/L (McKelvey et al., 2011; McRill et al., 2000; Sin & Tsang, 2003; Weldon et al., 2000). All of these studies used a reference value of 20 μg/L to organize biomarker values which was based on the Departments of Health in Hong Kong, New York State, Arizona, California, New Mexico and Texas, and the Centers for Disease Control and Prevention (CDC) and the U.S. Environmental Protection Agency (U.S. EPA). A total of 383 (67.3%) skin-lightening product users had urinary mercury levels above the reference values. Urinary mercury levels were assessed in 18 family members (i.e., husbands and children) of mercury-added skin-lightening product users and nine (50%) of their concentrations were above the reference value.

#### 3.5.2 Blood mercury levels

One study (Sin & Tsang, 2003) assessed total mercury levels in blood in addition to urinary mercury levels within 183 skin-lightening product users in Hong Kong. Blood mercury levels in users ranged from 0 to 82 μg/L with an overall median of 13 μg/L. This study used a reference value of 10 μg/L from the Department of Health in Hong Kong to organize biomarker values. A total of 119 (65%) users had blood mercury levels above the reference value.

#### 3.5.3 Hair mercury levels

Total hair mercury levels across 183 skin-lightening product users in Japan, Indonesia, Tanzania and Iran ranged between 0 and 900 μg/g (Abbas et al., 2020; Fakour & Esmaili-Sari, 2014; Harada et al., 2001; Kanwal et al., 2019; Kinabo, 2005). We excluded participants from Iran (n=36) since mercury concentrations were not available as discrete values (Fakour & Esmaili-Sari, 2014). We used hair mercury reference value ranges of below 1 μg/g, between 1 to 5 μg/g and above 5 μg/g adopted by Abbas et al. (2020) to organize biomarkers. Out of 147 users, a total of 103 (70.1%) individuals had hair mercury concentrations above 5 μg/g, 34 (23.1%) individuals were between 1 to 5 μg/g, and 10 (6.8%) individuals were below 1 μg/g. The overall pooled central median of hair mercury concentrations was 51.1 μg/g (IQR: 30.3-135.3) across 147 users (Abbas et al., 2020; Harada et al., 2001; Kanwal et al., 2019; Kinabo, 2005). Out of 63 non-users, 55 (87.3%) of individuals had hair mercury concentrations between 1 to 5 μg/g, 7 (11.1%) individuals were below 1 μg/g, and 1 (0.7%) individual was above 5 μg/g.

### 3.6 “Health impacts” Data Group

A total of 832 individuals from Kenya, USA, Jamaica and Hong Kong (Harada et al., 2001; McRill et al., 2000; Ricketts et al., 2020; Sin & Tsang, 2003; Weldon et al., 2000) self-reported symptoms associated with mercury poisoning. Nine individuals (81.8%) from Kenya (Harada et al., 2001) experienced tremors, lassitude, vertigo, neurasthenia and black and white blots reported after clinical examination through a questionnaire and undergoing a neurological test. In Jamaica, about 139 individuals reported experiencing itchiness (39.6%), irritability (38.1%), and “other” (headaches, depression and scars) (22.3%) (Ricketts et al., 2020). In individuals in the USA, and Hong Kong (McRill et al., 2000; Sin & Tsang, 2003; Weldon et al., 2000) the most reported outcomes included fatigue (n=245), nervousness/irritability (n=252), severe headaches (n=268), weakness (n=209), insomnia (n=189), anxiety/depression (n=167), and memory loss (n=155), tremors (n=123), and body/joint pain (n=97) (Figure 5). Although most participants in the USA and Hong Kong had elevated mercury levels in urine and blood that significantly exceeded reference values, 78% of participants in Hong Kong reported no symptoms. One study (McRill et al., 2000) offered clinical evaluations for affected individuals but doctors found no abnormalities related to health.

**Figure 5.**
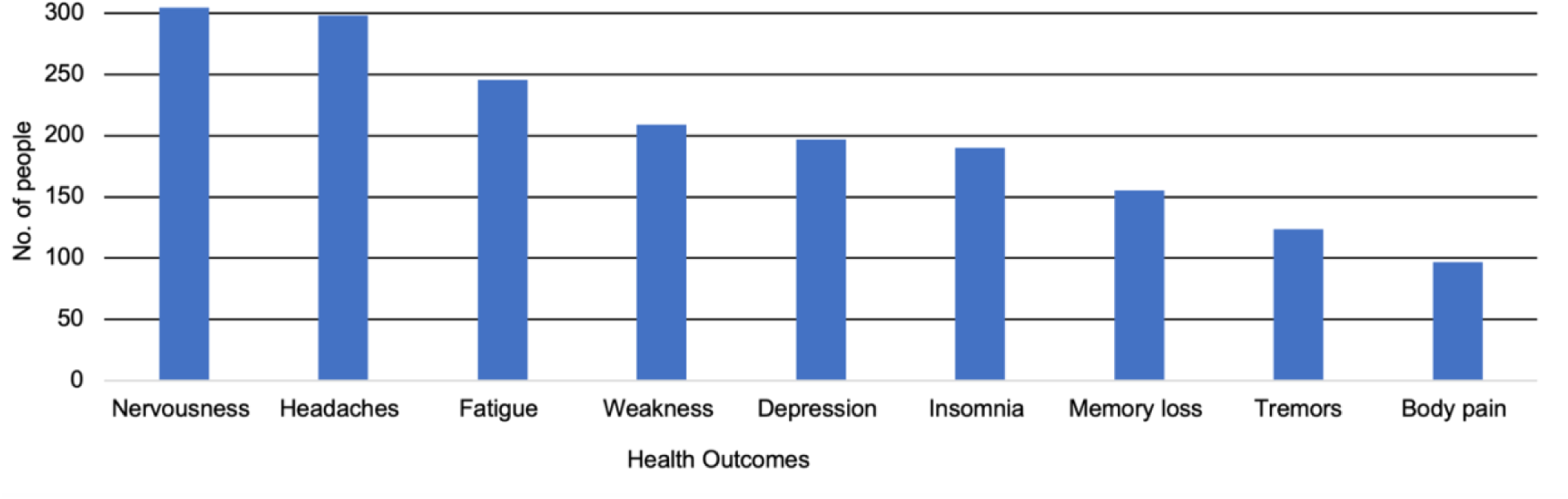
Count plot of self-reported health outcomes from 684 individuals from USA and Hong Kong measuring biomarker samples to determine presence of mercury poisoning and other associated health impacts from a targeted skin-lightening product (McRill et al., 2000; Sin & Tsang, 2003; Weldon et al., 2000).

## 4. Discussion

To our knowledge, this is the first systematic literature review characterizing the amount of mercury that human populations worldwide are being exposed to through the use of mercury-added skin-lightening products. In doing so, we conclude that mercury is still widely prevalent in skin-lightening products in many countries worldwide, and that relatively high exposures to mercury from skin-lightening products likely mediate adverse health outcomes among users and members of their household. Our conclusions are based on the compilation, synthesis and analysis of a dataset that consists of 787 skin-lightening products (manufactured in 33 countries and purchased from 20 countries worldwide), 1,042 mercury biomarker measurements taken from 863 individuals, usage patterns from 3,898 individuals, and self-reported health impacts from 832 individuals.

Mercury concentrations in skin-lightening products vary greatly among the products themselves, as well as among the studies and countries captured in this study. We are able to conclude this from our review of data from 787 skin-lightening products [(i.e., creams (70.1%), soaps (19.3%), facial cream (4.6%), and other products (6.0%)]. These products had mercury levels ranging from 0 to 314,387 μg/g with an overall pooled central median mercury concentration of 0.49 μg/g (IQR: 0.02-5.9 μg/g). Mercury concentrations in skin-lightening products varied across geographic regions with the highest median concentrations found in products purchased in the Eastern-Mediterranean, South-East Asian, and Western Pacific regions. Although we had a relatively large dataset to draw conclusions from, geographical data gaps were evident. Most notably is the relative absence of data from Europe, Southeast Asia, and many Latin American countries even though these are regions with populations known to use skin-lightening products (Sagoe et al., 2019).

A total of 19 out of 25 studies identified at least one skin-lightening product with mercury as an active ingredient (i.e., mercury concentration >1 μg/g). In addition to mercury, several studies have assessed other potentially toxic elements and chemical bleaching agents in skin-lightening products. Specifically, we note that 12 studies captured here evaluated only mercury, while 13 studies evaluated mercury in addition to other common active skin-lightening ingredients [e.g., hydroquinone, clobetasol propionate, kojic acid, corticosteroids (e.g., betamethasone, clobetasol propionate)] or trace elements [e.g., aluminum, arsenic, bismuth, cadmium, chromium, cobalt, copper, lead, iron, nickel, manganese, palladium, thallium, titanium, titanium dioxide, zinc] (Table S3). Some of these chemicals may be bioavailable, not easily metabolized by the body, and over time, cumulative exposures may contribute to a wide range of chronic health outcomes such as reproductive and developmental and neurological disorders, and renal damage (Bocca et al., 2014; Gbetoh & Amyot, 2016; Maneli et al., 2016).

In terms of the use of mercury-added skin-lightening products, the findings from the Sagoe et al. (2019) review paper showed that mercury is the second most popular bleaching agent globally after topical corticosteroids. Individuals aged 30 years or younger had the highest prevalence of using skin-lightening products (55.9%), followed by individuals aged 31 to 49 years (25.9%) and lastly individuals aged 50 years and above (6.1%). Based on their results they concluded that the use of skin-bleaching transcends an individual’s education level, relationship/marriage status, employment, and pregnancy stage (Sagoe et al., 2019). In our study, we found that there is great variability in skin-lightening product usage patterns across populations [i.e., application (i.e., face, whole body), frequency of application (per day), quantity used (g/month); and duration (months)]. Despite such variability, in general we conclude that most individuals who use these products apply skin-lightening items primarily on their face, typically from under 1 to 3 times per day, for less than a year, and use a quantity of 11 to 50 g per month. From this generalized conclusion, we can estimate mercury exposure for one year to be 150 μg mercury through the following calculation: (0.5 μg/g mercury, which was the overall pooled central median mercury concentration) * (25 g product per month) * 12 months. This estimate can significantly vary based on factors including product type and skin characteristics.

From the 2018 UN Global Mercury Assessment, it was determined that in background populations with insignificant exposures to mercury that individuals likely have mercury levels in blood, hair, and urine that are generally less than 5 μg/L, 2 μg/g, and 3 μg/L respectively (Basu et al., 2018). In the current study, we observe that most of the biomarker measures (i.e., median values) in users of skin-lightening products greatly exceeded the aforementioned levels found in background populations. We draw this conclusion from 1,042 biomarker measurements taken from 863 skin-lightening product users, family members and control groups. This is a relatively large mercury biomonitoring dataset though not one without challenges. Most of the data sets identified presented their biomarker data in relation to reference values set by their respective Departments of Health (McRill et al., 2000; Sin & Tsang, 2003; Weldon et al., 2000). While this made it challenging to make specific comparisons, we do note that the highest biomarker concentrations in these studies were well above their respective reference values. The studies with the largest sample sizes were case series studies of individuals with known exposure to different popular skin-lightening products in the U.S. (n=292 individuals from the U.S.-Mexican border region) (McRill et al., 2000; Weldon et al., 2000) and (n=286) users from Hong Kong (Sin & Tsang, 2003). In these cases, the studies were conducted after numerous incidents of adverse reactions associated with using these products were reported to health officials, and thus we conclude that these may represent worst case scenarios. Deriving a control population in these studies is particularly challenging though among users in US-Mexican border states (Arizona, California, New Mexico, and Texas), urinary mercury concentrations in 50% of immediate family members (i.e., husbands, children) were relatively high (29-340 μg/L) (McRill et al., 2000). This illustrates that mercury exposures among those who are in close proximity with users may also be elevated. Similar cases have been seen among pesticide applicators, where spouses and children of farmers had detectable levels of glyphosate in their urine without being present during the application process (Acquavella et al., 2004). Therefore, in addition to the users themselves, it is important to study family members and individuals who are in close proximity to skin-lightening product users to better understand their exposures and associated health risks.

Through our search, we flagged case reports in which health outcomes were observed in association with the use of mercury-added skin-lightening products (Table S4). Case reports are detailed medical reports that describe novel or unusual occurrence in patients and shared for scientific and educational purposes. Here, a total of 16 case report studies were compiled from seven countries (Barbados, China, USA, Belgium, UK, Philippines, and Mexico, Germany) with 37 individuals (users and non-users) between the ages of 10-months and 50 years old. The skin-lightening products mentioned had mercury concentrations between 1,762 and 56,000 μg/g mercury (versus 0.5 μg/g, which was the overall pooled central median mercury concentration we calculated here) or contained 0.07-6.5% mercury. Five individuals mentioned using products between six weeks to 10 years before reporting symptoms. The case reports showed that some individuals have very high levels of total mercury in urine (max=170 μg/L) and blood (max=2,620 μg/L), and inorganic mercury in hair (max=5,617 μg/g). Individuals in these cases studies presented signs of mercury intoxication, nephrotic syndrome, membranous glomerulonephritis, and hypertension. There were four reported cases of infants under the age of five that presented signs of mercury intoxication after being in contact with their mothers that were using mercury-added skin-lightening products (Copan et al., 2015; Kamani et al., 2019; Ori et al., 2018). The youngest reported case was a 10-month-old toddler who was admitted to the hospital after presenting symptoms including anorexia, hypertension, and regression in neuro-motor skills after his mother used a skin-lightening product locally made in Mexico containing 56,000 μg/g of mercury for six weeks prior to symptom onset (Kamani et al., 2019).

To characterize mercury exposures through the use of skin-lightening items, it is important to understand the trade of these products. Almost half (48.9%) of the products identified [i.e., cream (n=233), soap (n=109), facial cream (n=2), other (n=41)] did not indicate the country that they were manufactured in, and thus we could not identify where a large majority of skin-lightening products hail from. When the manufacturing location was noted, Europe, South-East Asia, and Africa were most prominent. Our review also noted countries in Europe (France, Italy, Spain, and the U.K.) as well as the U.S. that manufactured skin-lightening products with high concentrations of mercury. Regulating skin-lightening products in low-income countries is particularly challenging since these regions may not have regulations, and those that do are largely unenforced (Michalek, Benn, et al., 2019). In contrast, the European Union Rapid Alert System for dangerous non-food products (RAPEX) identified about 266 skin-lightening cosmetic import violations across Europe between 2005-2018, of which 15.4% contained mercury (Michalek, Liu, et al., 2019). From our findings, skin-lightening products containing mercury as an active ingredient (mercury concentration >1 μg/g) were mainly manufactured in the Dominican Republic, Mexico, Pakistan, and Thailand, similar to where the European Union RAPEX and the U.S. FDA have reported import violations. We found that the skin-lightening products were purchased mainly in Africa, the Eastern Mediterranean, the Americas and Western Pacific regions (according to WHO classifications), and that this is in line with where these products are most commonly used (Peregrino et al., 2011; Prevodnik et al., 2018; Sagoe et al., 2019). We found that the number of countries that manufacture products (n=33) is greater than the number of countries in which they were purchased (n=20), supporting the need for stronger enforcement of existing legislation and increased restrictions on mercury in skin-lightening products.

Despite performing a systematic review and compiling a relatively large dataset to draw conclusions from, there are some limitations of this work (from which recommendations are offered). Foremost is that the studies identified in this review tended to focus on geographic regions with known prevalence and concerns surrounding mercury-added skin-lightening products. Even in well-studied geographic regions, the sample sizes in individual works were relatively small. Further, all studies suffered from sampling bias (non-randomization) making it difficult to generalize. As a result, there is not enough representative data to conclude with high certainty the quantity of skin-lightening products containing mercury and the number of affected individuals worldwide. Nonetheless, our approach was able to derive a dataset from which we can suggest that mercury is still widely prevalent as an active ingredient (mercury concentration >1 μg/g) in many skin-lightening products worldwide, and that exposures among users is relatively high. These represent a good start and moving forward there is a need for more quality studies in this area.

Most individual biomarker concentrations in studies were either not stated or grouped together based on reference values set out by various agencies and health departments. Aside from the case series studies, biomarker studies generally sampled small groups of individuals likely due to the difficulty in isolating individual users and the negative stigma surrounding usage of skin-lightening products. There were very few studies collected that looked at control groups and non-users in the same areas or in areas where there are no suspected concerns. It can be difficult to isolate mercury exposures from the use of skin-lightening products, versus other notable sources (e.g., dental amalgams or seafood consumption). To help tease apart exposures to different chemical forms and sources of mercury, detailed survey instruments can be coupled with human biomarker measurements (Basu et al., 2018; UNEP/WHO, 2008). Mercury in hair generally reflects exposure to organic methylmercury (usually from the diet) though external contamination by mercury-added skin lightening products may complicate this measure and necessitate the use of advanced mercury speciation techniques. Mercury in urine reflects exposure to inorganic and elemental forms of mercury, and as a non-invasive and relatively inexpensive biomarker to sample this may be particularly attractive in exposure assessment studies (Basu et al., 2018). Finally, mercury in whole blood can provide information on recent exposures to both organic and inorganic mercury. In general, total mercury values are measured in hair, urine, or blood, though studies that employ chemical speciation or mercury stable isotope measures can glean data from which deeper understandings may be realized. With relatively simple training on sampling principles and quality assurance practices, human biomonitoring studies can be added to health studies in this area to help increase understanding of exposures.

All studies included in this review assessed skin-lightening products for the concentration of total mercury and assessed products using reliable instrumentation. The study quality of most studies was moderately ranked. We assessed study quality using the guidance from the framework published by Hu et al. (2018) and OHAT (2015) and found numerous issues in terms of exposure detection bias (e.g., need more information on quality control techniques including the use of reference materials and replicate measures especially for studies that sent samples to external laboratories for analysis), selection bias (e.g., need studies to carefully report on sampling techniques and breakdown of individuals concentrations of mercury in biomarker samples, and include more information on mercury exposure characteristics), and other biases (e.g., studies need to have more information on user demographics and concentrations of mercury that individuals are using to better understand mercury levels in biomarkers). As a result, we excluded 134 skin-lightening product samples from three studies (Hamann et al., 2014; Murphy et al., 2009; Ricketts et al., 2020) that used XRF analyzers and commercial screening kits. There are many concerns surrounding the validity and reliability of measurements taken from XRF instruments and screening kits since they do not have low detection limits or analytical reliability like CVAAS, ICP-MS or ICP-OES (McComb et al., 2014). Moving forward, we advocate that laboratories strengthen their capacity to test these products using higher quality measures (e.g., CVAAS, ICP-MS, AAS, PIXIE).

The Minamata Convention is driven by protecting human health (i.e., Articles 1, 16, 17, 19) and one means for increasing public information, awareness and education (Article 18) is to educate the public on the effects and exposure of mercury and mercury compounds on human health in collaboration with other relevant intergovernmental and non-governmental organizations (UNEP, 2019). Although the skin-lightening products industry generates billions of dollars annually, the products identified in this review represent a very small portion of the global inventory (Glenn, 2008). There is a strong need to create advocacy campaigns led by local health agencies and civil society to increase awareness and educate the community of the adverse health implications associated with skin-lightening, specifically skin-lightening products containing mercury. Advocacy campaigns run by national health authorities, governments and members of the public need to work together to educate users on the potential dangers of using mercury-added skin-lightening products. National programs to harmonize biomonitoring of exposed individuals need to be pursued in order to increase our understanding of human exposures. Governments need to create collaborative inventories of products with harmful levels of mercury, create legislation, and establish standards (e.g., contact information of manufacturer, country of manufacture, ingredient list) for imported products. There is a need for all retailers (e.g., stores, vendors, online) to be monitored by law enforcement and required to remove products that contain harmful levels of mercury.

## 5. Conclusion

The objective of this study was to increase our understanding of worldwide human mercury exposure and associated health risks from the use of skin-lightening products. To achieve this, we conducted a systematic review to collect and analyze data from 41 peer-reviewed papers, within which we further examined mercury concentrations in human biomarkers and skin-lightening product samples, along with usage patterns and associated health impacts. From this work, we can conclude that mercury widely exists as an active ingredient in skin-lightening products and that there is large variability in human exposures. While knowledge gaps and limitations exist (e.g., non-random selection bias, geographic regions with no data), these synthesized findings help increase our understanding of the health risks associated with the use of these products. In addition, we believe that the information in this study will be critical in helping regulatory agencies and inter-governmental organizations, particularly the Minamata Convention on Mercury and the WHO, better understand the global prevalence of mercury-added skin-lightening products over space and time.

## Supporting information

Bastiansz et al. Supplemental Excel File

Bastiansz et al. Supplemental Material

## Data Availability

All data produced in the present study are contained in the manuscript as well as within the supplemental material.

## ACKNOWLEDGEMENTS

We acknowledge funding support from the Canada Research Chairs Program and the PURE CREATE program funded by Natural Sciences and Engineering Research Council of Canada’s (NSERC). We thank Jenny Eng for technical and administrative support.

## DISCLAIMERS

The authors declare no competing financial interests.

