## Supplementary material for "Mercury exposure and health risks associated with use of skin-lightening products: A systematic review": Bastiansz et al. Supplemental Material

### Table of Contents

**Table S1.** Measurement instruments and abbreviations used for skin-lightening products analysis.

**Table S2.** Measurement instruments and abbreviations used for human biomarker analysis.

**Table S3.** Different toxic chemicals assessed in papers in the "Hg in products" data group.

**Table S4.** Case reports of individuals that experienced an adverse reaction following exposure to a mercury-added skin-lightening product.

**References Cited List**

**Additional File – "Supplemental Excel File" with:**
Excel Table S1. Database of mercury levels in skin-lightening products
Excel Table S2. Database of usage of skin-lightening products (application, frequency, duration, quantity)

**Table S1.** Measurement instruments and abbreviations used for skin-lightening product analysis.

| Measurement instrument used for testing skin-lightening cosmetics | No. of samples (%) |
|---|---|
| Atomic absorption spectroscopy (AAS) | 94 (11.94%) |
| Screen-printed silver electrode (AgSPE) | 3 (0.38%) |
| Cold vapor atomic absorption spectrometry (CVAAS) | 466 (59.21%) |
| Flame atomic absorption spectrometry (FAAS) | 6 (0.76%) |
| Inductively coupled plasma mass spectrometry (ICP-MS) | 38 (4.83%) |
| Inductively coupled plasma-optical emission spectrometry (ICP-OES) | 78 (9.91%) |
| Inductively coupled plasma atomic emission spectrometry (ICP-AES) | 49 (6.23%) |
| Particle Induced X-Ray Emission (PIXE) | 27 (3.43%) |
| Unknown | 26 (3.30%) |

**Table S2.** Measurement instruments and abbreviations used for human biomarker analysis.

| Measurement instrument used for testing human biomarkers | No. of samples (%) |
|---|---|
| Atomic absorption spectroscopy (AAS) | 40 (3.8%) |
| Advanced mercury analyzer (AMA) | 62 (6.0%) |
| Cold vapor atomic absorption spectrometry (CVAAS) | 11 (1.1%) |
| Inductively coupled plasma mass spectrometry (ICP-MS) | 13 (1.3%) |
| Particle Induced X-Ray Emission (PIXE) | 148 (3.4%) |
| Unknown | 768 (73.7%) |

**Table S3.** Different toxic chemicals assessed in papers in the "Hg in products" data group.

| No. | Reference | Toxic metals assessed |
|---|---|---|
| 1 | (Agorku et al., 2016) | Mercury, Hydroquinone |
| 2 | (Murphy et al., 2009) | Mercury |
| 3 | (Ho et al., 2017) | Mercury |
| 4 | (Zainy, 2015) | Mercury, Cadmium, Bismuth, Palladium, Arsenic, Titanium, Titanium dioxide, Thallium |
| 5 | (Harada et al., 2001) | Mercury |
| 6 | (Peregrino et al., 2011) | Mercury |
| 7 | (Gbetoh & Amyot, 2016) | Mercury, hydroquinone and clobetasol propionate |
| 8 | (Al-Saleh et al., 2012) | Mercury, Titanium dioxide, Hydroquinone, Corticosteroids, |
| 9 | (Yang et al., 2014) | Mercury |
| 10 | (Salama, 2016) | Lead, Mercury, Cadmium, Arsenic, Copper, Nickel, Cobalt, Manganese, Chromium, Aluminum |
| 11 | (Wang & Zhang, 2015) | Mercury |
| 12 | (McKelvey et al., 2011) | Mercury |
| 13 | (Ashraf et al., 2020) | Mercury, Cadmium, Lead, Arsenic, Chromium, Nickel, Cobalt, Copper, Zinc, Iron |
| 14 | (Selvaraju et al., 2020) | Mercury, Arsenic, Cadmium, Lead |
| 15 | (Abbas et al., 2020) | Mercury |
| 16 | (Kanwal et al., 2019) | Mercury |
| 17 | (Kinabo, 2009) | Mercury |
| 18 | (Voegborlo et al., 2008) | Mercury |
| 19 | (Alqadami et al., 2017) | Mercury, Arsenic, Bismuth, Cadmium, Lead, Titanium |
| 20 | (Alqadami et al., 2013) | Mercury, Arsenic, Bismuth, Cadmium, Lead, Titanium |
| 21 | (Maneli et al., 2016) | Mercury, Betamethasone, Clobetasol propionate, Hydroquinone; Kojic acid |
| 22 | (Ababneh & Al-Momani, 2018) | Mercury, Lead, Cadmium, Nickel |
| 23 | (Mohammed et al., 2017) | Mercury, Arsenic |
| 24 | (Amponsah et al., 2014) | Mercury |
| 25 | (Cristaudo et al., 2013) | Mercury, Cadmium, Cobalt, Chromium, Nickel, Lead |

**Table S4.** Case reports of individuals that experienced an adverse reaction following exposure to a mercury-added skin-lightening product.

| Authors, year | Ethnicity/ Race | Country | WHO Region | Clinical Details |
|---|---|---|---|---|
| (Drescher et al., 2013) | Barbadian | Barbados | PAHO | In 2010, high mercury exposure was discovered in four Barbadian women of African ancestry (aged 39-54) during a study analyzing mercury body-burden amongst individuals working in the fishing industry. All four women were using or had used skin bleaching creams, and had hair inorganic mercury concentrations 361-5617ug/g and spot urine inorganic mercury samples 7-135 ug/L. |
| (Soo et al., 2003) | Indonesian | China | WPRO | A 34-year-old Indonesian woman working as a domestic helper in Hong Kong for a year and had two weeks of frothy urine and ankle swelling. She had been using a skin whitening cream from Indonesia on her face every day for five years and had elevated blood mercury levels of 163nmol/L (59.09ug/L) and a 24-hour urinary mercury excretion rate of 754.6nmol/day, and the mercury concentration of the cream was 1762 ppm. |
| (Sun et al., 2017) | Chinese | China | WPRO | From 2009 to 2017, 16 female patients (aged 19-50 years) were diagnosed with chronic mercury intoxication at the First Affiliated Hospital of Zhengzhou University. All patients had a history of using skin lightening creams and one cream sample product contained 19,742 ± 379 ppm of mercury. The 24-hour urine mercury content of these patients was within 0.037-0.170mg/L (37ug/L to 170ug/L), and patients complained about body pain, tremors, and had differing levels of insomnia, irritability, and depression. |
| (Dickenson et al., 2013) | Mexican | USA | PAHO | During a research study looking at environmental chemical exposure in pregnant women and their infants, 1 participant out of 77 had elevated levels of mercury - whole blood mercury level: 15.16 ug/L, umbilical cord blood: 5.8 ug/L and urine mercury level 40ug/L. The participant had been using two skin lightening creams purchased in Mexico, which contained 21,000 and 30,000 ppm of mercury. |
| (Bwomda et al., 2005) | African | Belgium | EURO | A 24-year-old African woman presented at the emergency department six months after developing glucocorticosteroid induced arterial hypertension, weight gain, amenorrhea, and a deep cutaneous ulcer on the right foot, swollen erythematous right leg, and thin skin with deep red striae over her abdomen, and signs of Cushing's syndrome. She admitted to using skin bleaching cream for at least 10 years and applied over full body. |
| (Chakera et al., 2011) | Pakistani | United Kingdom | EURO | Two cases of Pakistani women (aged 44 and 26 years), they were referred for investigation of the nephrotic syndrome, renal biopsies showed that they had membranous glomerulonephritis associated to skin lightening creams. Their serum and urinary mercury-creatinine ratio was 150nmol/L and 16.5nnmol/mmol, and 233 nmol/l and 77.5nmol/mmol, respectively. |

| | | | | |
|---|---|---|---|---|
| (Ori et al., 2018) | Mexican? | USA | PAHO | A 17-month-old toddler, had 3 weeks of fussiness, constipation, and decreased appetite, developed a limp with tenderness in the right knee, whole blood mercury, 26 ug/L, and random spot mercury level of 243 ug/g creatinine. Mother and grandmother had been using skin lightening creams, neither of them complained about symptoms but had elevated first void urine mercury levels of 197ug/g creatinine (mother) and 222 ug/g creatinine (grandmother), and cream contained mercury between 27,000 and 34,000 ug/g. |
| (Copan et al., 2015) | Mexican? | USA | PAHO | Three cases between 2010-2014 of a 20-month-old, and a 39-year-old and her 4-year-old child. The three individuals had elevated mercury levels between 52 ug/g to 482ug/g creatinine, all associated with creams that were between 20,000-57,000 ug/g. |
| (Zhang et al., 2014) | Chinese | China | WPRO | A 28-year-old woman patient was admitted to the hospital in January 2010, after complaining of pain in both limbs and facial edema for two weeks. She was diagnosed with nephrotic syndrome. The patient used a homemade skin lightening cream for 11 months that contained 6.80 ug/g, and her blood and urinary mercury concentration was 220 umol/L and 469 umol/L, respectively. |
| (Tlacuilo-Parra et al., 2001) | Mexican? | Mexico | PAHO | A 30-year-old woman in 1996 experienced burning face pain and excessive perspiration after using a daily facial skin lightening cleanser for five years with listed calomel. Her urine mercury was 150ug/L and whole blood mercury was 30 ug/L. |
| (Tang et al., 2006) Tang 2006 | Chinese | Hong Kong | WPRO | A 34-year-old Chinese woman developed nephrotic syndrome, minimal change disease and increased mercury concentrations in blood (24.8 µg/L) and urine (57.4 µg/L) after using a skin-lightening cream contain 30,000 ug/g of mercury daily for 4 months. Her proteinuria returned to normal after 9 months, and her blood and urine became normal after 1 and 9 months respectively after cessation of usage. |
| Choi 2002 | Chinese | Hong Kong | WPRO | A 32-year-old woman developed nephrotic syndrome, minimal change nephropathy and a raised blood mercury concentration (48.6 µg/L) after using a cosmetic cream (3.4% Mercury) for 3 months. After penicillamine therapy, her blood mercury concentration dropped to 10.8 µg/L, and her renal function returned to normal within 3 months. The skin-lightening cream she used was found to contain 0.07 – 0.21% mercury. |
| (Chan et al., 2001) | Chinese | Hong Kong | WPRO | A 38-year-old woman had an asymptomatic increase in blood mercury concentration (18.8µ g/L) and a urine mercury excretion (69 µg/day) after daily use of a cosmetic cream for several months. The cream contained 6.5% mercury. |
| (Barit et al., 2020) | Filipino | Philippines | WPRO | A 27-year-old Filipino female presented pruritic erythematous papules and plaques on the bilateral axillae after using a whitening cream containing 6400 ppm of mercury. Positive reactions to mercury ammonium chloride and thiomersal were shown in patch test results. |

| | | | | |
|---|---|---|---|---|
| (Kuehn, 2020) | Mexican American | USA | PAHO | According to a CDC report, a 47-year-old Mexican American female developed dysesthesias and upper extremity weakness, and later hospitalized for two weeks with slurred speech, blurry vision, an unsteady gait after using two skin-lightening creams on her face twice daily for 7 years. She had blood mercury levels of 2620 μg/L and urinary mercury levels of 110 μg/L and underwent chelation therapy. Skin-lightening creams were purchased in Mexico and contained 12,000 ppm of mercury; further testing of creams detected methyl mercury iodide. |
| (Kamani et al., 2019) | Mexican American? | USA | PAHO | A ten-month-old male was admitted to hospital with a one-month history of a progressive desquamative papular rash, anorexia, weight loss, hypertension, and regression in neuro-motor milestones. The patient had whole blood mercury levels at 251 nmol/L and underwent empiric succimer chelation therapy for three weeks. The patient's mother purchased a locally made skin cream in Mexico containing elemental mercury at 56,000 ppm and had been using it on her face/neck twice daily for 6 weeks before onset of patient's symptoms. |
